# Different mutations in SARS-CoV-2 associate with severe and mild outcome

**DOI:** 10.1101/2020.10.16.20213710

**Authors:** Ádám Nagy, Sándor Pongor, Balázs Győrffy

## Abstract

**Introduction:** Genomic alterations in a viral genome can lead to either better or worse outcome and identifying these mutations is of utmost importance. Here, we correlated protein-level mutations in the SARS-CoV-2 virus to clinical outcome.

**Methods:** Mutations in viral sequences from the GISAID virus repository were evaluated by using “hCoV-19/Wuhan/WIV04/2019” as the reference. Patient outcomes were classified as mild disease, hospitalization and severe disease (death or documented treatment in an intensive-care unit). Chi-square test was applied to examine the association between each mutation and patient outcome. False discovery rate was computed to correct for multiple hypothesis testing and results passing a FDR cutoff of 5% were accepted as significant.

**Result:** Mutations were mapped to amino acid changes for 2,120 non-silent mutations. Mutations correlated to mild outcome were located in the ORF8, NSP6, ORF3a, NSP4, and in the nucleocapsid phosphoprotein N. Mutations associated with inferior outcome were located in the surface (S) glycoprotein, in the RNA dependent RNA polymerase, in the 3’-to5’ exonuclease, in ORF3a, NSP2 and N. Mutations leading to severe outcome with low prevalence were found in the surface (S) glycoprotein and in NSP7. Five out of 17 of the most significant mutations mapped onto a 10 amino acid long phosphorylated stretch of N indicating that in spite of obvious sampling restrictions the approach can find functionally relevant sites in the viral genome.

**Conclusions:** We demonstrate that mutations in the viral genes may have a direct correlation to clinical outcome. Our results help to quickly identify SARS-CoV-2 infections harboring mutations related to severe outcome.

## INTRODUCTION

There are seven human coronaviruses including MERS, Human-HKU-1, Human NL63, Human 229E, Human OC43, SARS-CoV, and SARS-CoV-2. The natural host of this latest RNA virus is the Chinese rufous horseshoe bat (*Rhinolophus sinicus*) and the transfer to human initiated the ongoing COVID-19 outbreak at the end of 2019 ^1^. The mortality rate of SARS-CoV-2 in the overall population is low ^2^, and the infections have at most limited impact on the total number of respiratory-virus associated mortality ^3^. However, when the virus strikes a critically ill patient, mortality can go up to 26% ^4^.

The linear genome of the SARS-CoV-2 virus has 29,903 bases and harbors 25 genes ^5^, the reference sequence in accessible in GeneBank using the accession number MN908947. Phylogenetic analysis of SARS-CoV-2 genomes show three variants termed A, B and C which have different distribution when comparing sequences from Asia, Europe or the Americans ^6^. The viral genes include among other an envelope protein, an RNA dependent RNA polymerase, a surface glycoprotein, an exonuclease, a methyltransferase, and 11 nonstructural proteins. Some of these are within the virus, but others, including the spike glycoprotein, the membrane glycoprotein, and the envelope protein are on the viral surface.

In theory, any functional or structural viral gene can have an effect on the efficiency of a virus and both mutations ^7^ or alteration in the expression ^8^ can increase pathogenicity. It is important to emphasize that even the untranslated regions of a coronavirus can have important role in the viral replication as has been previously demonstrated for the 3’ untranslated region ^9^. SARS-CoV-2 is no different compared to other viruses and new mutations continually pop up with the spread ^10^. Some mutations uncovered in the SARS-CoV-2 virus lead to a novel RNA-dependent-RNA polymerase variant ^11^, while other genomic changes drive the evolution and the spread of the virus by resulting in a more transmissible form of the virus ^12^. Mutations potentially making the virus more transmissible have a significant evolutionary advantage as has been demonstrated for the SARS-CoV-2 variant with spike G614 which mainly replaced D614 between February and July 2020 ^13^.

In this context, the most important question is to identify viral mutations leading to different patient outcomes. Mutations resulting in a mild disease could facilitate the spread of the virus and thereby maintain the outbreak. Other mutations leading to a more severe disease need immediate attention to prevent detrimental outcomes. Here, our goal was to identify and rank mutations associated with altered patient outcome by simultaneously correlating outcomes to all mutations across a large cohort of patients.

## MATERIALS AND METHODS

### Data source

All available SARS-CoV-2 (taxid: 2697049) viral nucleic acid sequences were downloaded from the GISAID virus repository (https://www.gisaid.org/). The sequences were acquired in FASTA format. Those viral sequences were selected where the entire viral nucleic acid sequence was published. A second filtering was executed to include only virus genomes with available patient follow-up status.

### Mapping of mutations to viral genes

The mutations were evaluated using the CoVsurver (https://corona.bii.a-star.edu.sg). To achieve this, the viral sequences in .FASTA format were used as the query and the “hCoV-19/Wuhan/WIV04/2019” was used as the reference. The analysis was run by using batches of 1000 samples in one run. Protein mutations don’t have overlaps, and the genomic boundaries of the various proteins in the WIV04 reference genome are displayed in **Table 1**.

**Table 1.**
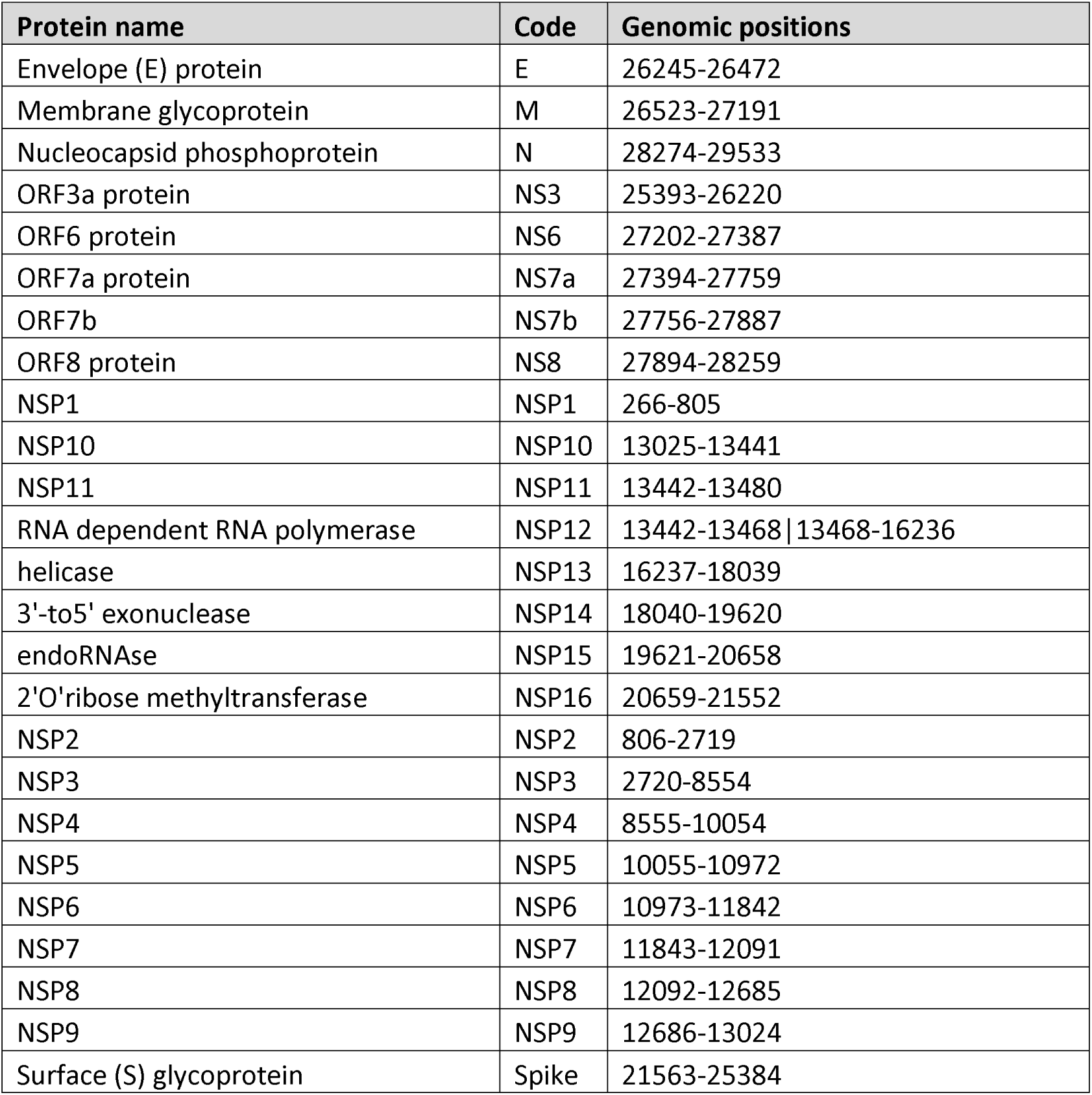
Genomic boundaries of the SARS-CoV-2 proteins in the WIV04 reference genome

| Protein name | Code | Genomic positions |
| --- | --- | --- |
| Envelope (E) protein | E | 26245-26472 |
| Membrane glycoprotein | M | 26523-27191 |
| Nucleocapsid phosphoprotein | N | 28274-29533 |
| ORF3a protein | NS3 | 25393-26220 |
| ORF6 protein | NS6 | 27202-27387 |
| ORF7a protein | NS7a | 27394-27759 |
| ORF7b | NS7b | 27756-27887 |
| ORF8 protein | NS8 | 27894-28259 |
| NSP1 | NSP1 | 266-805 |
| NSP10 | NSP10 | 13025-13441 |
| NSP11 | NSP11 | 13442-13480 |
| RNA dependent RNA polymerase | NSP12 | 13442-13468 \| 13468-16236 |
| helicase | NSP13 | 16237-18039 |
| 3'-to5' exonuclease | NSP14 | 18040-19620 |
| endoRNAse | NSP15 | 19621-20658 |
| 2'O'ribose methyltransferase | NSP16 | 20659-21552 |
| NSP2 | NSP2 | 806-2719 |
| NSP3 | NSP3 | 2720-8554 |
| NSP4 | NSP4 | 8555-10054 |
| NSP5 | NSP5 | 10055-10972 |
| NSP6 | NSP6 | 10973-11842 |
| NSP7 | NSP7 | 11843-12091 |
| NSP8 | NSP8 | 12092-12685 |
| NSP9 | NSP9 | 12686-13024 |
| Surface (S) glycoprotein | Spike | 21563-25384 |

### Clinical classification

As the patient samples were annotated with all together more than sixty different outcome classification, we had to coerce these into three major categories.

Patients who were “asymptomatic”, were “not hospitalized”, had a “mild” disease, were at “home” were all assigned to have a “mild” disease. Also patients who were treated at outpatient departments, were quarantined or were treated by the physician network were classified as “mild”.

Patients who definitely needed medical care were assigned to the “hospitalized” group. These include those “hospitalized”, “inpatient”, “discharged”, “released”, and “recovered”. In addition, combinations of the annotations which included any of these were also assigned into this cohort (e.g. “initially hospitalized” or “to be hospitalized”).

Finally, patients with detrimental outcome were allocated to the “severe” cohort. These include those “deceased”, those with a “severe” disease, those who entered “intensive care units”. Also any combination of these with other annotations (e.g. “hospitalized / ICU”) were also added to this category.

### Statistical computation

All data processing and statistical analysis steps were performed in the R statistical environment v 3.6.3. Data processing was performed on 28^th^ July 2020. Chi-square test was applied to examine the association between each mutation and patient status data. False discovery rate using the Benjamini-Hochberg method was computed to correct for multiple hypothesis testing and only results passing a FDR cutoff of 5% were accepted as significant.

## RESULTS

### Dataset

All together 73,020 SARS-CoV-2 viral nucleic acid sequences were available, and 72,331 of these included the entire viral nucleic acid sequence. Clinical data was available for 5,094 patients, and 3,184 of these had also follow-up data. This is a small fraction of the total data which implies that our findings could contain a sampling bias.

When looking on the clinical parameters of these patients, 55.6% were male and 38.2% were female (remaining samples did not had this information). The geographical origin of the samples cover the entire globe: 4.8% were from Africa, 45.4% from Asia, 9.4% from Central America, 26.7% from Europe, 6.4% from North America and 6.6% from South America. Collection of the samples happened between 30.12.2019 and 4.7.2020. Of all patients with a follow-up 625 had a mild disease, 2,341 had to be hospitalized and 218 patients had a severe disease.

### Mutation rate

All together 2,121 different mutations affecting the protein structure were identified, and 463 of these mutations were not present in samples with clinical follow-up. When looking on all mutations, we have identified on average 2.81 mutations in each sample.

As an internal control to validate any potential bias in the mutations prevalence related to patient proportions we computed the average sample numbers for each clinical outcome cohort, and these overlapped with the actual clinical outcome (wild type and mild outcome mean = 623, wild type and hospitalized mean = 2,336, wild type and severe outcome mean = 217).

When analyzing the correlation to clinical outcome across all mutations, 141 mutations reached statistical significance at FDR<5%. The complete list of these mutations with sample numbers in each cohort is displayed in **Supplemental Table 1** and patient level mutation data is provided in **Supplemental Table 2**.

### Mutations related to mild disease

In order to concentrate only on mutations with a clinical relevance, we selected only those mutations which were present in at least 2% of the samples (this corresponds to a cutoff of at least 64 patient samples with a mutation). When looking on mutation related to mild outcome, only six mutations passed all filtering criteria - L84S in the ORF8 protein, L37F in the NSP6 protein, G196V in the ORF3a protein, F308Y in the NSP4 protein, and the S197L and S202N mutations in the nucleocapsid phosphoprotein. The complete list as well as distribution among patient samples is provided in **Table 2**.

**Table 2.** SARS-CoV-2 mutations correlated to mild outcome in 3,184 patients with available genomic and follow-up information were found in five distinct genes.

| Protein name | Protein mutation | N wild type + mild outcome | N wild type + hospitalized | N wild type + severe outcome | N mutant + mild outcome | N mutant + hospitalized | N mutant + severe outcome | Chi squared test p value |
| --- | --- | --- | --- | --- | --- | --- | --- | --- |
| ORF8 protein | L84S | 394 | 2153 | 206 | **231** | 188 | 12 | **4.0E-80** |
| NSP6 | L37F | 500 | 2094 | 210 | **125** | 247 | 8 | **4.0E-13** |
| ORF3a protein | G196V | 450 | 2297 | 211 | **175** | 44 | 7 | **1.0E-112** |
| NSP4 | F308Y | 449 | 2294 | 211 | **176** | 47 | 7 | **2.9E-111** |
| Nucleocapsid phosphoprotein | S197L | 450 | 2294 | 211 | **175** | 47 | 7 | **2.4E-110** |
| Nucleocapsid phosphoprotein | S202N | 600 | 2297 | 218 | **25** | 44 | 0 | **4.0E-04** |

### Mutations associated with severe disease

When searching for mutations related to hospitalization or to severe outcome, we used the above filter of including only mutations present in at least 2% of the samples. All together nine mutations passed these criteria. These originated in seven genes: D614G and L54F in the surface (S) glycoprotein, P323L in the RNA dependent RNA polymerase, Q57H in the ORF3a protein, S194L, R203K, and G204R mutations in the nucleocapsid phosphoprotein, L177F in the 3’-to5’ exonuclease, and Q496P in the NSP2 protein.

In order not to miss mutation leading to deadly outcome we also included all mutations which were present in at least 10 patients with severe outcome. This additional analysis delivered two further mutations, the V1176F in the surface (S) glycoprotein and the L71F mutation in the NSP7 gene. These resulted in 27 and 11 severe outcomes after being spotted in 28 and 11 patients, respectively.

Interestingly, the overall prevalence of mutations leading to mild outcome (n=1,565) was smaller than the prevalence of those leading to worse outcome (n=6,696). Nevertheless, a significant proportion of the mutations (n=5,053) were not significantly correlated to any clinical outcome.

The complete list of all mutations correlated to severe disease is presented in **Table 3**.

**Table 3.** SARS-CoV-2 mutations correlated to hospitalization and severe outcome in 3,184 patients with available genomic and follow-up information were found in eight distinct genes.

| Protein name | Protein mutation | N wild type + mild outcome | N wild type + hospitalized | N wild type + severe outcome | N mutant + mild outcome | N mutant + hospitalized | N mutant + severe outcome | Chi squared test p value |
| --- | --- | --- | --- | --- | --- | --- | --- | --- |
| Surface (S) glycoprotein | D614G | 366 | 641 | 31 | 259 | 1700 | **187** | **6.2E-56** |
| RNA dependent RNA polymerase | P323L | 373 | 679 | 37 | 252 | 1662 | **181** | **3.2E-52** |
| ORF3a protein | Q57H | 527 | 1666 | 150 | 98 | **675** | **68** | **7.4E-11** |
| Nucleocapsid phosphoprotein | R203K | 551 | 1886 | 160 | 74 | **455** | **58** | **4.3E-07** |
| Nucleocapsid phosphoprotein | G204R | 552 | 1889 | 161 | 73 | 452 | **57** | **5.3E-07** |
| Surface (S) glycoprotein | V1176F | 625 | 2340 | 191 | 0 | 1 | **27** | **6.6E-78** |
| Nucleocapsid phosphoprotein | S194L | 624 | 2182 | 200 | 1 | 159 | **18** | **2.4E-10** |
| NSP7 | L71F | 625 | 2341 | 207 | 0 | 0 | **11** | **2.4E-33** |
| NSP2 | Q496P | 623 | 2280 | 211 | 2 | **61** | 7 | **1.4E-03** |
| 3'-to5' exonuclease | L177F | 624 | 2276 | 212 | 1 | **65** | 6 | **4.2E-04** |
| Surface (S) glycoprotein | L54F | 623 | 2262 | 213 | 2 | **79** | 5 | **1.5E-04** |

## DISCUSSION

We have simultaneously analyzed the correlation between patient outcome and all identified mutations resulting in amino acid sequence changes in the viral proteins. Strikingly, we have not only found a significant number of mutations, but some of these were correlated to mild diseases while other had a significant correlation to severe outcome.

Nucleocapsid phosphoprotein, the protein with most significant mutations was linked to both mild and severe patient outcome. All these changes are at a close genomic positions, S197L and S202N resulting in mild outcome and R203K, G204R, and S194L resulting in inferior outcome. Interestingly, when comparing the S197L (76% of mild outcome) to the S197L (less than 1% chance of a mild outcome) variants, the relative risk was extremely high. Interestingly, the majority of the nucleocapside phosphoprotein mutations mapped to a small stretch of amino acids from position 194 to 204. This region coincides with the phosphorylated “RS-motif” ^14^ which maps onto the intrinsically unstructured serine rich region 181-213 of the protein ^15^. Phosphorylation of this site is known to play important roles such as recruitment of host RNA helicase DDX1 which facilitates template readthrough and enables longer subgenomic mRNA synthesis (https://www.uniprot.org/uniprot/P59595). This observation needs further follow-up – especially because the nucleocapsid phosphoprotein is one of the potential drug targets against SARS-CoV-2 ^16^.

Overall, we have observed more mutations in the structural proteins (spike and nucleocapsid phosphoprotein) than in non-structural proteins. Of note, destabilization mutations in nonstructural proteins were suggested to represent a potential mechanism differentiating SARS-CoV-2 from SARS ^17^.

Previously, a set of common deletions were identified in the spike protein of SARS-CoV-2 ^18^. Other deletions were also validated by RT-PCR ^19^. However, due to the scarce number of identical changes we could not evaluate a potential link between deletions and patient outcome.

Importantly, our findings might contain a sampling bias, since only a fraction of the available genomes had patient outcome data. On the other hand, five out of 17 potentially significant mutations (listed in Tables 2 and 3) map to an about 10 amino acid long, functionally important region of the nucleocapsid phosphoprotein which leads us to believe that the current statistical approach can reveal functionally important sites within the COVID 19 genome.

Coronaviruses have generally a stable genome which changes very little over time ^20^. A fundamental question of SARS-CoV-2 research is whether or not the virus can get weaker or stronger with time. Our findings suggest that there are mutations that can support either of these changes so the theoretical possibility is there that in the future the viral effect will shift towards milder or more severe patient outcomes.

## Data Availability

All available SARS-CoV-2 (taxid: 2697049) viral nucleic acid sequences were downloaded from the GISAID virus repository (https://www.gisaid.org/).
The mutations were evaluated using the CoVsurver (https://corona.bii.a-star.edu.sg).

https://corona.bii.a-star.edu.sg

## ACKNOWLEDGEMENTS

The research was financed by the 2018-2.1.17-TET-KR-00001 and KH-129581 grants and by the Higher Education Institutional Excellence Programme of the Ministry for Innovation and Technology (MIT) in Hungary, within the framework of the Bionic thematic programme of the Semmelweis University as well as by OTKA grant 12065 provided by MIT, Hungary to Pázmány University. The authors wish to acknowledge the support of ELIXIR Hungary (www.elixir-hungary.org) as well as the advice of Drs Sebastian Maurer-Stroh (Bioinformatics Institute, A*STAR, Singapore) and Balázs Ligeti (Pázmány University, Budapest).

## REFERENCES

1 Andersen, K. G., Rambaut, A., Lipkin, W. I., Holmes, E. C. & Garry, R. F. The proximal origin of SARS-CoV-2. Nat Med 26, 450–452, doi:10.1038/s41591-020-0820-9 (2020).

2 Roussel, Y. et al. SARS-CoV-2: fear versus data. Int J Antimicrob Agents 55, 105947, doi:10.1016/j.ijantimicag.2020.105947 (2020).

3 Giraud-Gatineau, A. et al. Comparison of mortality associated with respiratory viral infections between December 2019 and March 2020 with that of the previous year in Southeastern France. Int J Infect Dis 96, 154–156, doi:10.1016/j.ijid.2020.05.001 (2020).

4 Grasselli, G. et al. Baseline Characteristics and Outcomes of 1591 Patients Infected With SARS-CoV-2 Admitted to ICUs of the Lombardy Region, Italy. JJAMA, doi:10.1001/jama.2020.5394 (2020).

5 Wu, F. et al. A new coronavirus associated with human respiratory disease in China. Nature 579, 265–269, doi:10.1038/s41586-020-2008-3 (2020).

6 Forster, P., Forster, L., Renfrew, C. & Forster, M. Phylogenetic network analysis of SARS-CoV-2 genomes. Proc Natl Acad Sci U S A 117, 9241–9243, doi:10.1073/pnas.2004999117 (2020).

7 Chapman, S., Hills, G., Watts, J. & Baulcombe, D. Mutational analysis of the coat protein gene of potato virus X: effects on virion morphology and viral pathogenicity. Virology 191, 223–230, doi:10.1016/0042-6822(92)90183-p (1992).

8 Tober, C. et al. Expression of measles virus V protein is associated with pathogenicity and control of viral RNA synthesis.J Virol 72, 8124–8132, doi:10.1128/JVI.72.10.8124-8132.1998 (1998).

9 Williams, G. D., Chang, R. Y. & Brian, D. A. A phylogenetically conserved hairpin-type 3’ untranslated region pseudoknot functions in coronavirus RNA replication.J Virol 73, 8349-8355, doi:10.1128/JVI.73.10.8349-8355.1999 (1999).

10 van Dorp, L. et al. Emergence of genomic diversity and recurrent mutations in SARS-CoV-2. Infect Genet Evol 83, 104351, doi:10.1016/j.meegid.2020.104351 (2020).

11 Pachetti, M. et al. Emerging SARS-CoV-2 mutation hot spots include a novel RNA-dependent-RNA polymerase variant. J Transl Med 18, 179, doi:10.1186/s12967-020-02344-6 (2020).

12 Yurkovetskiy, L. et al. SARS-CoV-2 Spike protein variant D614G increases infectivity and retains sensitivity to antibodies that target the receptor binding domain. bioRxiv, doi:10.1101/2020.07.04.187757 (2020).

13 Korber, B. et al. Tracking Changes in SARS-CoV-2 Spike: Evidence that D614G Increases Infectivity of the COVID-19 Virus. Cell, doi:10.1016/j.cell.2020.06.043 (2020).

14 Peng, T. Y., Lee, K. R. & Tarn, W. Y. Phosphorylation of the arginine/serine dipeptide-rich motif of the severe acute respiratory syndrome coronavirus nucleocapsid protein modulates its multimerization, translation inhibitory activity and cellular localization. JFEBS J 275, 4152–4163, doi:10.1111/j.1742-4658.2008.06564.x (2008).

15 Dosztanyi, Z., Csizmok, V., Tompa, P. & Simon, I. IUPred: web server for the prediction of intrinsically unstructured regions of proteins based on estimated energy content. Bioinformatics 21, 3433–3434, doi:10.1093/bioinformatics/bti541 (2005).

16 Yadav, R., Imran, M., Dhamija, P., Suchal, K. & Handu, S. Virtual screening and dynamics of potential inhibitors targeting RNA binding domain of nucleocapsid phosphoprotein from SARS-CoV-2. Journal of biomolecular structure & dynamics, 1–16, doi:10.1080/07391102.2020.1778536 (2020).

17 Angeletti, S. et al. COVID-2019: The role of the nsp2 and nsp3 in its pathogenesis. J Med Virol 92, 584–588, doi:10.1002/jmv.25719 (2020).

18 Liu, Z. et al. Identification of common deletions in the spike protein of SARS-CoV-2. J Virol, doi:10.1128/JVI.00790-20 (2020).

19 Holland, L. A. et al. An 81-Nucleotide Deletion in SARS-CoV-2 ORF7a Identified from Sentinel Surveillance in Arizona (January to March 2020). J Virol 94, doi:10.1128/JVI.00711-20 (2020).

20 Vijgen, L., Lemey, P., Keyaerts, E. & Van Ranst, M. Genetic variability of human respiratory coronavirus OC43. Journal of virology 79, 3223-3224; author reply 3224-3225, doi:10.1128/JVI.79.5.3223-3225.2005 (2005).

